# Measles Burden Estimation Using Local Gaussian Process Classifiers

**DOI:** 10.1101/2022.05.17.22275226

**Authors:** Xiaoxiao Li, Zhou Lan, Matthew Ferrari, Murali Haran

## Abstract

Measles is a highly contagious viral disease and remains a severe public health problem. Monitoring measles cases provides a powerful tool to identify outbreaks and epidemics. However, burden estimates of measles are challenging to obtain because of heterogeneous surveillance systems and a lack of resources for rapid laboratory tests means that many cases are reported based on symptoms alone, which has low specificity. We consider diagnostically confirmed measles case data in Ethiopia between 2009 and 2017 and propose a local Gaussian process binary classifier with spatial dependence to provide case predictions for untested individuals based on age, vaccination status, and location. By applying our modeling framework to untested suspected reported cases, we provide more accurate burden estimates at the district level. We validate our methods through simulation studies. We also find that our approach, which provides burden estimates at the district level, highlights temporal variation in the specificity of symptom-based diagnosis.

## 1 Introduction

Measles is a highly contagious viral disease that can cause significant morbidity and mortality, especially in children. In the pre-vaccination era, measles caused an estimated 2.6 million deaths each year (WHO, 2019) and is still one of the leading causes of child morbidity and mortality in many African countries with high birth rate (Cutts et al., 2021). Though the widespread use of measles vaccines has greatly decreased the incidence of measles infection and mortality rate (Portnoy et al., 2019), it remains a serious public health concern.

Reliable and robust surveillance is a critical tool in the design and implementation of vaccination, control, and elimination policies (Cutts et al., 2021; Beyene et al., 2016). The World Health Organization (WHO) has been conducting case-based surveillance for measles in 44 countries out of 47 WHO member states on the WHO African Region and the performance varies widely by country (Masresha et al., 2018). WHO member states submit information on suspected measles cases, clinically defined as a fever and a maculopapular (non-vesicular) rash and at least one of cough, coryza, or conjunctivitis, as part of routine surveillance activities. Given the ubiquity of these symptoms, especially in children, the clinical case definition has low positive predictive value, particularly in settings with low prevalence (Hutchins et al., 2004), and laboratory confirmation is an important component of measles surveillance. Laboratory confirmation of suspected measles cases is commonly done via enzyme-linked immunosorbent assay (ELISA) for measles-specific immunoglobulin M (IgM) antibodies. However, due to lack of medical infrastructure and professionals, a significant proportion of suspected measles cases reported (fever and rash) are not confirmed with a laboratory test in many low- and middle-income settings (Masresha et al., 2018). This partial observation and confirmation relative to the true incidence adds uncertainty to measles burden estimation over time, especially in countries with poor vital registration and disease surveillance systems.

Estimates of the burden of measles disease and mortality, as well as evaluation of the performance of vaccination programs, rely on statistical models fit to reported time series of measles cases (Simons et al., 2012; Eilertson et al., 2019; Dixon et al., 2021). The low specificity of the measles clinical case definition (Hutchins et al., 2004) implies that an uncertain fraction of these reported cases – determined by the rate of diagnostic confirmation, the background prevalence of measles, and the prevalence of non-measles febrile rash – could lead to biases in these critical operational metrics. Here we propose a hybrid approach to estimate the burden of reported measles cases by combining the diagnostically confirmed cases with an estimate of the number of untested cases that would be diagnostically confirmed, if tested.

In order to estimate the burden of cases among untested suspected cases, we study multiple classifiers: (i) a local logistic regression (Cockcroft et al., 2009) that builds separate classifier for each subdivision, (ii) a flexible Gaussian process-based version (cf. Williams and Rasmussen, 2006) of the local logistic regression, and (iii) a novel spatially varying local Gaussian process classifier that uses local (subdivision-level) information as well as information from neighboring regions. Introducing spatial dependence allows classifiers to borrow information from neighbors to better train a predictive model for districts with few or even no test confirmed data on record.

In this paper, we focus on measles burden estimation with partial surveillance information. Our contributions may be summarized as follows: (1) We propose a local Gaussian process binary classifier with spatial dependence to provide individual level test predictions using age, vaccination status and location information. (2) Our modeling framework provides more accurate burden estimation at a high spatial resolution, for example district level in Ethiopia. These burden estimates can help with analyses of the dynamics of the measles epidemic and provide guidance in policy design and operation.

The rest of the paper is organized as follows. In Section 2 we introduce the measles surveillance data in Ethiopia which motivated the development of our model. In Section 3 we present the structure of our model together with two other models that are closely related in our logistic model framework. In Section 4 we study our approach through extensive simulation both in classification and burden estimation. In Section 5 we show the results and findings of applying our models to Ethiopia surveillance data. In Section 6 we discuss the range of applications for our model and some directions for future study and research.

## 2 Measles Surveillance Data in Ethiopia (2009-2017)

Ethiopia is located in the Horn of Africa where measles remains endemic. Though there are efforts to achieve elimination goals, periodic measles outbreaks are still putting millions of people’s welfare at risk. To reduce and prevent the illness and death caused by measles, Ethiopia has been engaged in immunization plans among children in different ages since 1980 (Akalu, 2015) and initiated measles case based surveillance since 2003, which is supported by partial laboratory confirmation of suspected cases starting from 2004 (Federal Ministry of Health of Ethiopia, 2017, 2011). In this study, we extract the individual-level surveillance data in Ethiopia from 2009 to 2017 coordinated by the Ethiopian Public Health Institute (EPHI) which has two components: 13,409 cases who seek healthcare and have laboratory IgM status tested on record, and 31,853 cases who are suspected measles cases with febrile rash that are submitted by healthcare providers without IgM test status. Age (in months), vaccination status (in number of doses) and location information up to district level of each case are also being recorded. ^1^

Measles vaccination coverage has significant spatial dependence in Ethiopia (Takahashi et al., 2017; Geremew et al., 2019). For instance urban children are more likely to have received all eight basic vaccinations than rural children (65% vs. 35%), and the coverage is lowest in Affar (15%) and highest in Addis Ababa (89%) (CSA and ICF, 2017). As shown in Figure 1 (b), eastern provinces Afar, Somali have lowest vaccination coverage; the provinces to the west have higher vaccination coverage. The age distribution of all the cases reported also has variability on map of Ethiopia. As shown in Figure 1 (a), the mean age is higher in northern provinces such as Tigray, Afar, Amhara, and lower in the western provinces. Figure 1 (c) shows the number of reported cases at district level which also varies in space; we noticed some districts don’t have any cases reported in 2009-2017, 8 out of 70 districts reported cases less than 20, 5 among which have fewer than 10 cases; 6 of the districts have all cases reported in one measles status (all test positive or all negative). Lack of sufficient confirmed cases and imbalance among classes at these districts make it challenging to provide individual-level predictions and hence burden estimation.

**Figure 1:**
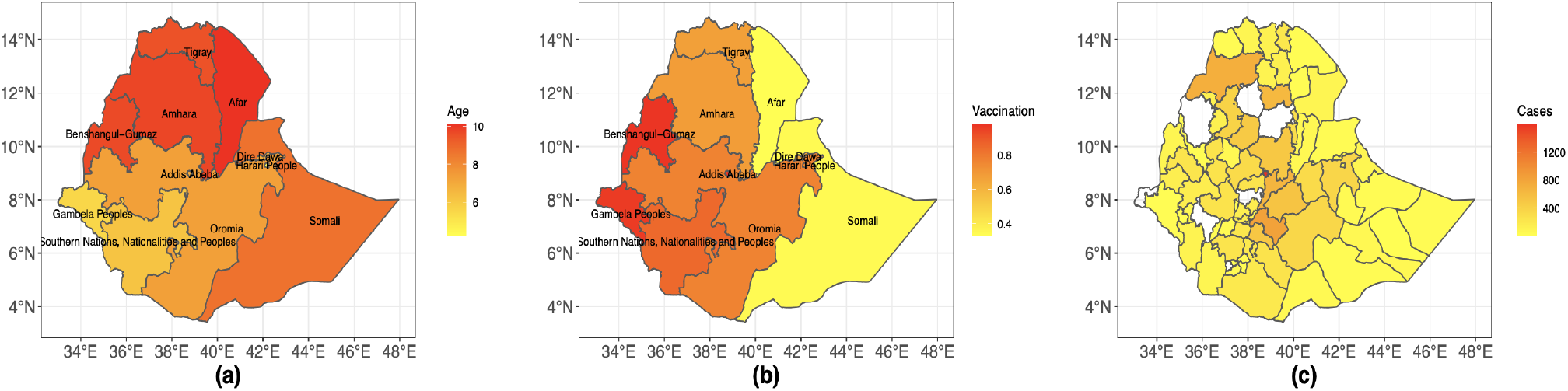
Distributions of (a) mean age at province level (b) mean number of vaccination received at province level and (c) number of cases at district level of all records with IgM test status on map of Ethiopia

Figure 2 is the time series plot that compares the number of laboratory confirmed cases and the reported suspected cases from 2009 to 2017. We notice that only a small proportion of cases received laboratory test results; confirmed cases have lower variability and suspected cases have relatively large variance over time. Though the trend between the two are almost the same, the magnitude of the ratio between the number of confirmed and suspected cases is not constant overtime. It indicates that with the purpose of providing accurate measles burden estimation based on the current surveillance data, purely relying on the suspected cases tends to overestimate the disease burden, whereas only using the confirmed cases will underestimate the true disease burden.

**Figure 2:**
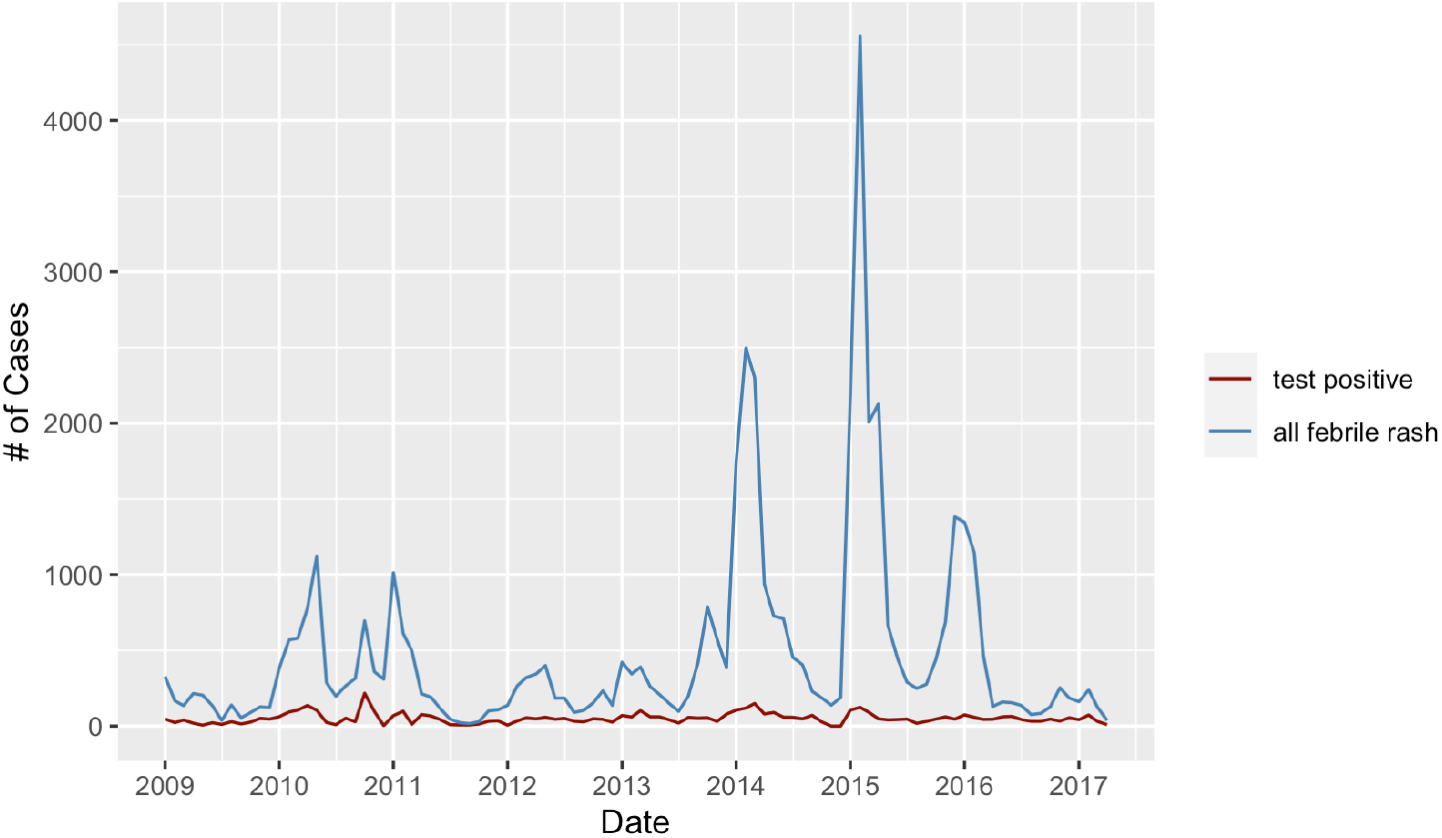
The aggregated number of confirmed cases and suspected cases from 2009 to 2017 in Ethiopia

## 3 Methods

Motivated by Ethiopia’s measles surveillance data, we present our proposed classification methods. This method provides individual-level prediction on measles test status (positive or negative) which can be further extended to the reported suspected cases with no laboratory confirmation to provide burden estimates that account for spatial heterogeneity. Classification methods like logistic regression, decision trees, Naive Bayes, support-vector machines and neural network (Hastie et al., 2009) have been widely applied in disease prediction (cf. Wiemken and Kelley, 2019). We consider various models based upon a flexible logistic regression framework. To build our classifier, we use training data consist of individuals with igM test for measles. There are multiple individuals observed at each of several locations across Ethiopia. We can then use our classifier to make predictions on the suspected cases that have not been tested but for whom we have vaccination status and age. Let *Y*_*i*_(*s*) ∈ {0, 1} be the measles status of the *i*th individual at location *s* where 0 and 1 respectively indicate a negative and positive test for measles. There are *J*_*s*_ individuals at the *s*th location, where *s* ∈ *𝒟* ⊆ R2, with *𝒟* representing the study region. Here *D* is Ethiopia while each *s* is the centroid of a district in Ethiopia. ***X***_*i*_(*s*) is a vector of *p* covariates for the *i*-th subject at location *s*, here this consists of vaccination status in doses (0-3) and age in month centered to zero. We provide our final adjusted measles burden estimates of people who visited the clinics by adding our inferred number of predicted measles cases from the reported suspected cases to the confirmed cases with laboratory results. In this section we outline three different classifiers for burden estimation: local logistic regression (LLR), spatial logistic regression (SLR), and varying coefficient spatial logistic regression (V-SLR).

### 3.1 Local Logistic Regression (LLR)

Logistic regression assumes that the response *Y*_*i*_(*s*) of subject *i* at location *s* follows a Bernoulli distribution with the probability of success *P*_*i*_(*s*) ∈ [0, 1], denoted as *Y*_*i*_(*s*) ∼ Bernoulli [*P*_*i*_(*s*)]. The probability of success *P*_*i*_(*s*) is then transformed into *μ*_*i*_(*s*) ∈ R via the logit function, i.e., 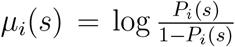 and thus 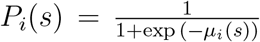. We model the transformed mean *μ*_*i*_(*s*) = ***X***_*i*_(*s*)***β*** + *β*_0_, where ***β*** is a vector of coefficients for their corresponding predictors and *β*_0_ is the intercept. Epidemiological studies have indicated that many covariate effects are region specific (Geremew et al., 2019; Geweniger and Abbas, 2020; Tesfa et al., 2022) and could be caused by factors such as access to health facilities, variations in healthcare operations and regional movements and contact patterns. We start with a baseline model that is a local logistic regression (LLR) fit independently to each subregion (provinces in the Ethiopian measles data).

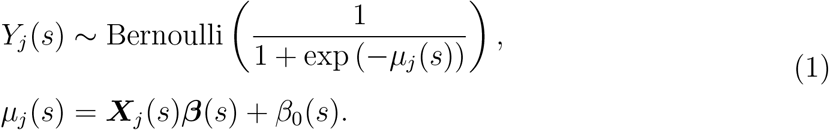

LLR essentially provides a logistic regression classifier for each location where ***β***(*s*) and *β*_0_(*s*) captures the heterogeneity in space of covariates and baseline risk. As shown in Figure 3, we map the coefficients of covariates fitted in each district and find that the covariate effects are spatially-varying with a gradient from southeast to northwest, which suggests that the model might benefit from assuming spatial dependence among the covariates; this acts as our motivation for the spatially varying coefficient model in Section 3.3.

**Figure 3:**
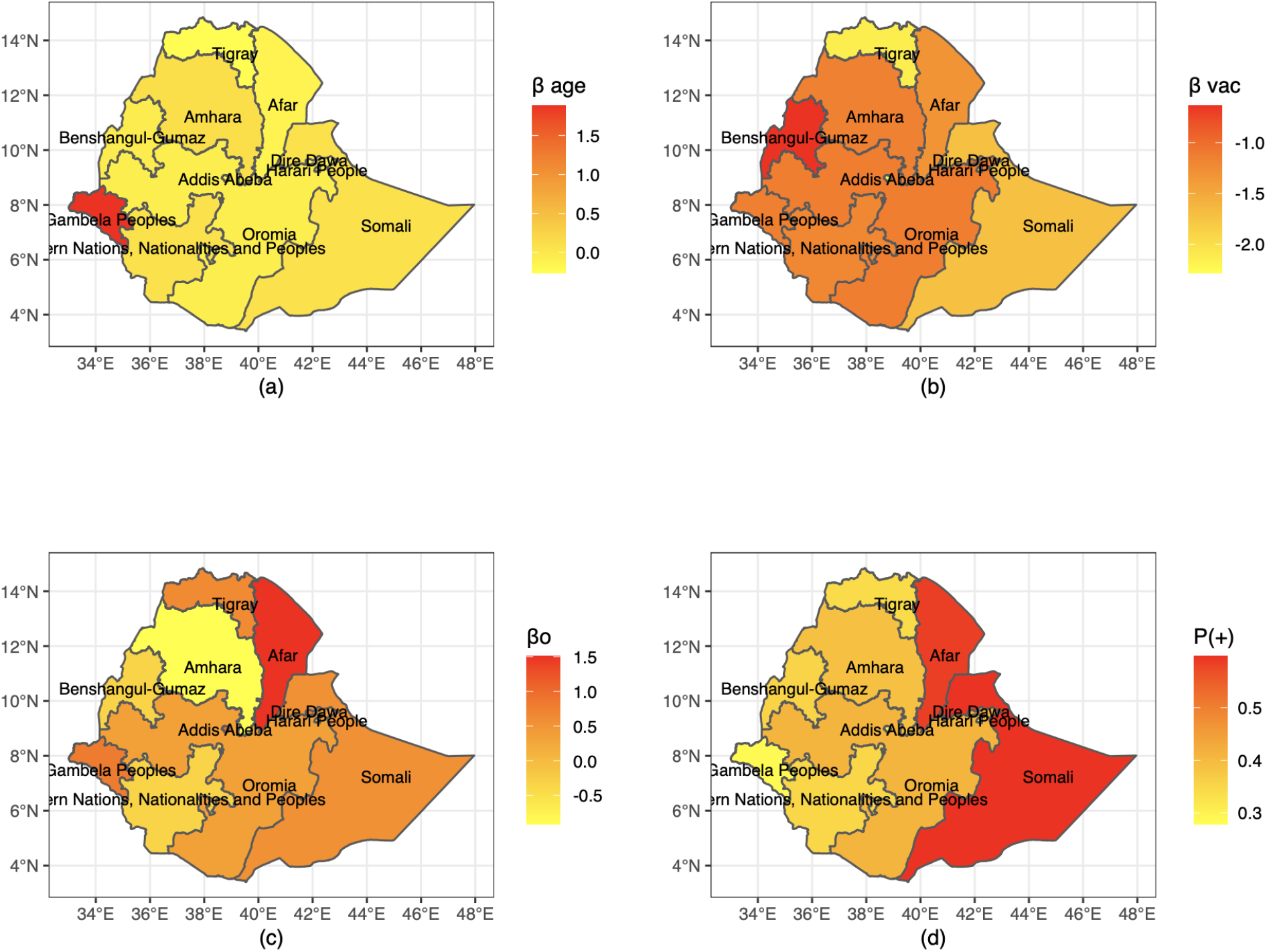
(1) Intercepts from LLR models (2) Coefficients of age from LLR models (3) Coefficient of vaccination status of LLR models (4) Mean positive rate on map of Ethiopia at province-level.

### 3.2 Spatial Logistic Regression (SLR)

LLR fails to describe the spatial dependence among districts. An approach to allowing for spatial dependence is to add spatially dependent random effects to the model, which replaces *β*_0_(*s*) in LLR with *W* (*s*) below. This class of models, commonly called spatial generalized linear models, is well studied in the literature on non-Gaussian spatial data (cf. Besag et al., 1991; Zhang, 2002; Haran, 2011; Hughes and Haran, 2013). The spatially dependent logistic regression model (SLR) is specified as follows: hierarchical model can be expressed as:

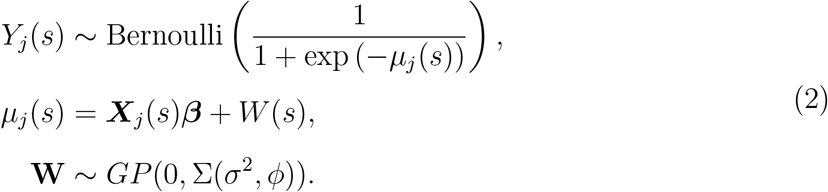

We model {*W* (*s*), *s* ∈ *D*} as a Gaussian process with mean 0 and an exponential covariance function to describe the spatial dependence among different locations: Σ^*i,j*^ = *Cov*(*W* (*s*_*i*_), *W* (*s*_*j*_)) = *σ*^2^ exp(−*d*_*i,j*_*/ϕ*), where *d*_*i,j*_ is the distance between location *i* and *j, σ*^2^ is the variance and *ϕ* is the spatial scaling parameter (larger value of *ϕ* has higher covariance). For any collection of *n* locations this implies that ***W*** = [*W* (*s*_1_), …, *W* (*s*_*n*_)] is an *n*−dimensional multivariate normal with an *n × n* covariance matrix. This model is also studied as Gaussian process binary classifier (Williams and Rasmussen, 2006) and can be intuitively understood as an extension of logistic regression which captures both the linear effect specified by predictors and the non-linear effects which are caused by the joint variability between districts described by the spatial dependence.

### 3.3 Varying Coefficient Spatial Logistic Regression (V-SLR)

LLR (Equation 1) and SLR (Equation 2) provide binary classification while allowing for spatial heterogeneity. However, as is the case for our Ethiopia data, it is common for many locations to have a limited number of observations or even zero observations. This poses challenges for training a reliable classifier locally; in some cases the few observations may all belong to a single class (all positive or all negative), making the problem even more challenging. SLR model handles these situations better than LLR model but only utilizes neighboring information through the spatial dependence in random effect *W* (*s*). In addition, we find that the coefficients for our Gaussian process classifier also vary smoothly in space (see Figure 3). We propose a novel model that utilizes this information based on SLR and further borrow information from neighboring locations through the coefficients of covariates which can be achieved by modeling each predictor ***β***_*k*_ = [*β*_*k*_(*s*_1_), …, *β*_*k*_(*s*_*n*_)] for *k* = {1, …, *p*} as a Gaussian process with mean 0 and covariance Σ_*k*_ where Σ_*k*_ is a *n* by *n* matrix. Together with the random effect ***W*** ∼ *GP* (0, Σ_*W*_) where Σ_*W*_ is also *n* by *n* in dimension, we utilize *p* + 1 Gaussian processes to describe the spatial dependence among classifiers in different locations. The full hierarchical model can be expressed as:

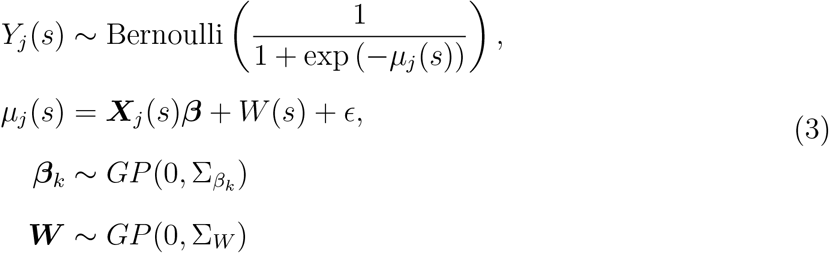

where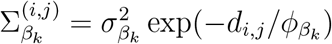 and 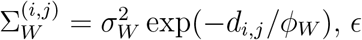, *ϵ* is added into the model to account for the imbalance in the response variable. We refer to Equation 3 as varying coefficient spatial logistic regression model (V-SLR). V-SLR meets our expectation in the exploratory analysis in Figure 3 where spatial variation presents in both coefficients and random effects, and can overcome the drawbacks encountered by LLR and SLR by constructing a classifier for locations with few or even zero observations based on the spatial dependency with the neighboring locations.

Details regarding prior distributions and the Markov chain Monte Carlo (MCMC) algorithm used to carry out Bayesian inference are provided in the Supplement. For efficiency and extendability, we implement the MCMC algorithms using the language Nimble (de Valpine et al., 2017).

## 4 Simulation Studies

In this section, we use synthetic data under multiple experimental settings to evaluate the performance of our proposed V-SLR method, together with LLR and SLR as two baseline models. With the purpose of providing classification of measles status at individual-level at various districts, we evaluate the performance of the classifiers in terms of true positive rate (TPR), true negative rate (TNR), accuracy (ACC) and area under the receiver operating characteristic curve (AUC). We also evaluate the performance of burden estimation both at country-level and district-level. In both simulation studies, we use the administrative map of Ethiopia with province and district levels.

### 4.1 Performance on Classification

In order to conduct simulations based on synthetic data that resembles real data in Ethiopia, we keep the individual demographic records on age, vaccination status and location information the same as the surveillance data in Ethiopia. However, we generate the measles status for each individual according to the logistic model with spatial dependency as described in Section 3.3. This provides control over the ground truth of the model through pre-specified coefficients and random effects. We use these covariates as predictors and an exponential covariance function to describe spatial dependence^2^, Σ^(*i,j*)^ = *σ*^2^ exp(−*d*_*i,j*_*/ϕ*), with parameters *σ*^2^, *ϕ >* 0. The coefficients for age, vaccination status are generated by *β*_*k*_ ∼ *GP* (0, Σ_*k*_) where *k* ∈ {age, vaccination}, and random effects *W* generated by *W* ∼ *GP* (0, Σ_*W*_). The variance parameters for Σ_*k*_ and Σ_*W*_ are set to be 1. We design three levels of spatial dependence on the covariates and random effects by varying *ϕ*: low (*ϕ* = 200), medium (*ϕ* = 500), and high (*ϕ* = 800) spatial dependence. We set *ϵ* to 0 for all the three panels of synthetic data.

All the simulation studies were conducted through 10-fold cross-validation. We randomly split the data into 10 folds of similar size; for each fold, we train the 3 classifiers using the remaining folds as training data set and evaluate the performance of these classifiers on the hold out fold as testing set. We report the average of metrics in the simulation study across the 10 folds in Table 1. As shown in the table, the overall performance of SLR and our proposed V-SLR model are both better than LLR model on all the 3 levels of spatial dependence in the simulation setting, which implies that we can improve the performance of individual-level classifiers through introducing spatial dependence. In the medium and high spatial dependence settings, V-SLR are in general performing better than that of SLR. For the evaluation metrics, true positive rate, also referred to as sensitivity, is evaluating the probability that an actual positive will test positive. True negative rate refers to the probability of a negative test, given that one does not have measles. Accuracy gives the overall prediction accuracy regardless of positive or negative cases; area under curve in our study refers to the integration of the area under the operating characteristic curve, which plots the true positive rate against the false positive rate at varying discrimination threshold. For TPR and TNR, V-SLR model outperforms LLR model, but shows no obvious advancement over SLR. On metric of ACC and AUC, V-SLR model outperforms LLR model and SLR model in both medium and high spatial dependence settings. Our heuristic to choose between the three models involves analyzing the correlogram of the intercepts and coefficients fitted from LLR model. We used the ‘correlog’ function in the ‘ncf’ package (Ottar and Cai, 2016) in the statistical language R to draw correlogram plots. If we see significant spatial dependence in the intercept term (if the confidence band excludes 0), then spatial dependence models (SLR and V-SLR models) are preferred; if further spatial dependence presents in coefficients term, then the V-SLR model is preferred. The SLR model and V-SLR models are more computationally expensive to fit than the LLR model due to the additional spatial dependence term and the fact that only LLR is easily parallelized.

**Table 1:** Performance of LLR, SLR and V-SLR models under simulation settings with 3 levels of spatial dependence. Monte Carlo error reported in parentheses; bold numbers represent the best performance (only if the improvement is statistically significant).

| Spatial<br>Dependence | Evaluation<br>Metric | LLR | SLR | V-SLR |
| --- | --- | --- | --- | --- |
| low ($\phi = 200$) | TPR | 0.67 (0.03) | **0.86** (0.03) | 0.79 (0.02) |
|  | TNR | 0.69 (0.02) | **0.85** (0.04) | 0.77 (0.03) |
|  | ACC | 0.67 (0.01) | **0.86** (0.01) | 0.78 (0.01) |
|  | AUC | 0.73 (0.02) | **0.93** (0.01) | 0.83 (0.01) |
| medium ($\phi = 500$) | TPR | 0.59 (0.01) | 0.83 (0.02) | 0.82 (0.02) |
|  | TNR | 0.80 (0.05) | 0.82 (0.02) | **0.85** (0.01) |
|  | ACC | 0.65 (0.01) | 0.82 (0.01) | **0.84** (0.01) |
|  | AUC | 0.70 (0.02) | 0.89 (0.02) | **0.93**(0.02) |
| high ($\phi = 800$) | TPR | 0.61 (0.01) | 0.82 (0.01) | **0.85** (0.01) |
|  | TNR | 0.65 (0.03) | 0.83 (0.02) | 0.86 (0.03) |
|  | ACC | 0.63 (0.01) | 0.83 (0.01) | **0.85** (0.01) |
|  | AUC | 0.70 (0.02) | 0.89 (0.02) | **0.93** (0.02) |

**Table 2:** 10-fold cross-validation performance comparing scaling up method vs LLR, SLR and V-SLR models on percentage of burden estimation CIs that covers the ground truth at district-level. Monte Carlo error reported in parentheses.

| Spatial<br>Dependence | Scale Up | LLR | SLR | V-SLR |
| --- | --- | --- | --- | --- |
| Low ($\phi = 200$) | 78.3% (1.9%) | 81.0% (3.4%) | 96.6% (2.0%) | 97.0% (2.1%) |
| Medium ($\phi = 500$) | 82.3% (3.6%) | 85.4% (3.1%) | 97.7% (1.7%) | 98.3% (1.5%) |
| High ($\phi = 800$) | 85.9% (2.9%) | 91.3% (3.0%) | 97.9% (2.0%) | 97.4% (1.5%) |

### 4.2 Performance on Burden Estimation

After obtaining the trained classifiers using the three models, LLR, SLR and V-SLR, we utilize them to estimate the disease burden on testing data set. A burden estimation method is desirable if it gives accurate estimates while keeping a relatively smaller uncertainty; what’s more, in our study, we are interested in better estimates of disease burden not only on country level, but also on more granulated level in district. Therefore, we first provide the burden estimates and 95% confidence intervals (CI) from models on country level, and then compare on whether the constructed CIs cover the ground truth of disease burden. In our analysis at country level, all CIs constructed from the three models cover the ground truth with comparable uncertainty. Then we break down the burden estimate into district and compare confident intervals constructed through LLR model, SLR model, V-SLR model and a scaling up method. The scaling up method is a common way of providing burden estimates which first obtains a point estimate of test positive rate at each district based on training data and then scale it up to the testing data with uncertainty derived from Binomial distribution. In the cross-validation, we construct district-level CIs for each method and record the mean number of districts which has disease burden successfully covered across the folds. In Table 1, we report the performance of district level coverage of CIs in percentage in 3 different spatial dependence levels. LLR, SLR and V-SLR all outperforms the scale up methods, which shows the importance of introducing statistical models in disease burden estimation at finer geographical levels. SLR and V-SLR both outperforms LLR shows that by introducing spatial dependence in the model, the performance of burden estimation at the district level improves, which is valuable for formulating targeted policy. V-SLR covers more ground truth measles burden at district-level at low and medium spatial dependence settings, but not significantly better.

## 5 Measles Burden Estimation in Ethiopia

In this section we apply our methods to measles surveillance data set from Ethiopia. As in the simulation study, we first compare the performance of classifiers at individual-level, then obtain country-level and district-level disease burden estimates from these classifiers. In exploratory data analysis, we observe the distribution of vaccination doses are highly concentrated at 0 and 1, with a smaller proportion on 2 and 3 doses. The phenomenon is partially attributed to the mistakes in the process of case recording, for example, some doctors may treated this question as whether or not the case have received measles vaccination before or the individual forgot the accurate number of doses received, thus some cases who received 2 or 3 doses are recorded as 1. As a result, we coded our vaccination status as binary, with 0 represents no vaccination received before and 1 as have received vaccination regardless the number of doses; the binary coding of vaccination also helps with interpretability on the coefficients. In addition, we observe a nonlinear relationship between age and proportion of measles test positive, which indicates adding a quadratic term of age in the classifiers as predictors. Note that in the original data, there were some mismatches in the location information at district and province level, for example, multiple objects in district Mirab Gojjam are matched to province Amhara and it actually in province Oromia. We made corrections to 5 districts in total with such mismatches.

### 5.1 Performance on Classification

We first investigate the performance of classifiers introduced in Section 3 (LLR, SLR and V-SLR models) in the application to the individual-level binary classification of measles status in Ethiopia. All the data analysis results are conducted through 5-fold cross-validation in order to evaluate the performance among different classifiers. Performance between different classifiers is evaluated in terms of TPR, TNR, ACC and AUC as in Section 4.1 and we report the average of each metric across the 5 folds. SLR and V-SLR models are classifiers at district-level, LLR is fitted at province-level because of insufficient valid observations at multiple districts.

Table 3 provides the cross-validation performance of the three models where V-SLR model outperforms LLR and SLR models on all the 4 metrics, which indicates the importance of including spatial dependence in improving predictability of individual-level measles status. LLR is fast in computation and the results are not far off from SLR and V-SLR model.

**Table 3:** 5-fold cross-validation performance of LLR, SLR and V-SLR models in terms of TPR, TNR, ACC and AUC. (Monte Carlo standard error less than or equal to 0.01)

| Evaluation<br>Metric | LLR | SLR | V-SLR |
| --- | --- | --- | --- |
| TPR | 0.62 | 0.63 | **0.64** |
| TNR | 0.75 | 0.74 | **0.75** |
| ACC | 0.70 | 0.70 | **0.72** |
| AUC | 0.74 | 0.74 | **0.76** |

The quadratic term of age has coefficient 0.004 which is shared across all district; the spatial varying coefficient of age in Figure 4 (a) has both positive and negative values in districts. Intuitively, as age goes up, the immunity gained either from vaccination or infection should accumulate and thus lower the risk of getting measles. This is true among children of various ages, however, due to the difference in behavior, the number of adult observation is a lot fewer and the risk is higher among these adults who visit clinics since they tends not to visit hospital unless the phenomenon is severe and treatment is needed. Figure 4 (b) shows the coefficients on vaccination on district level of V-SLR model. The coefficients are negative for all districts which corresponds to the scientific fact that vaccination reduces the risk of getting infected with measles. The plot also indicates that the impact of vaccination has spatial heterogeneity across districts. Figure 4 (c) is the plot of the random effects, which can be viewed as the background risk of being measles positive consisting all possible factors in addition to age and vaccination status.

**Figure 4:**
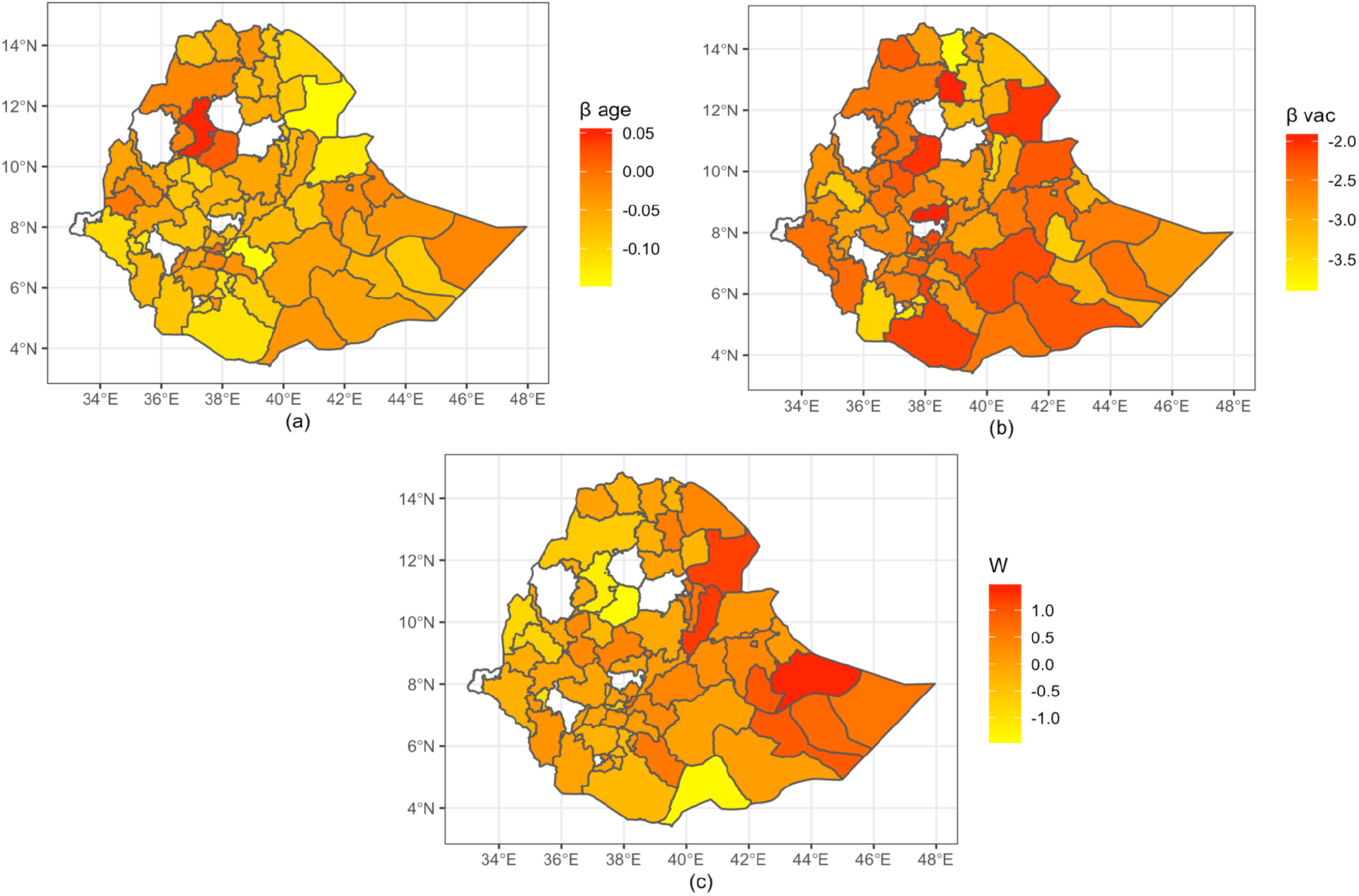
(a) Coefficients of age (b) Coefficients of vaccination (c) Random effects in V-SLR model on map of Ethiopia.

### 5.2 Performance on Burden Estimation

In the measles surveillance data of Ethiopia, we constructed individual-level measles status classifiers based on age, vaccination status and location information, and can utilize them to provide burden estimation on all the reported suspected cases without laboratory confirmation. The constructed confidence intervals at country level are comparable and they all captured the ground truth burden of measles: the simple scaling up method (from Section 4.2) (1220, 1339), LLR (1228,1328) SLR (1230,1333) and V-SLR (1223, 1324).

When we break down the burden estimation to district level, the confidence intervals for each district behaves differently for different methods. To better compare and demonstrate the results, we pick one fold of the cross-validation study and centralize the 95% CIs by subtracting their ground truth, i.e., the horizontal line at 0 represents ground truth of burden after center correcting at that district. As a result, a CI that covers the true disease burden at a district will contain 0. Figure 5 shows whether, at the district level, each CI from the 4 methods covers the ground truth disease burden or not. The CIs are displayed vertically by methods with blue represents ground truth being covered and red represents not being covered. CIs constructed through V-SLR model covers the ground truth disease burden at most districts, followed by SLR and LLR model; the scaling up method (introduced in Section 4.2), though widely used, has the least accurate CIs at the district level.

**Figure 5:**
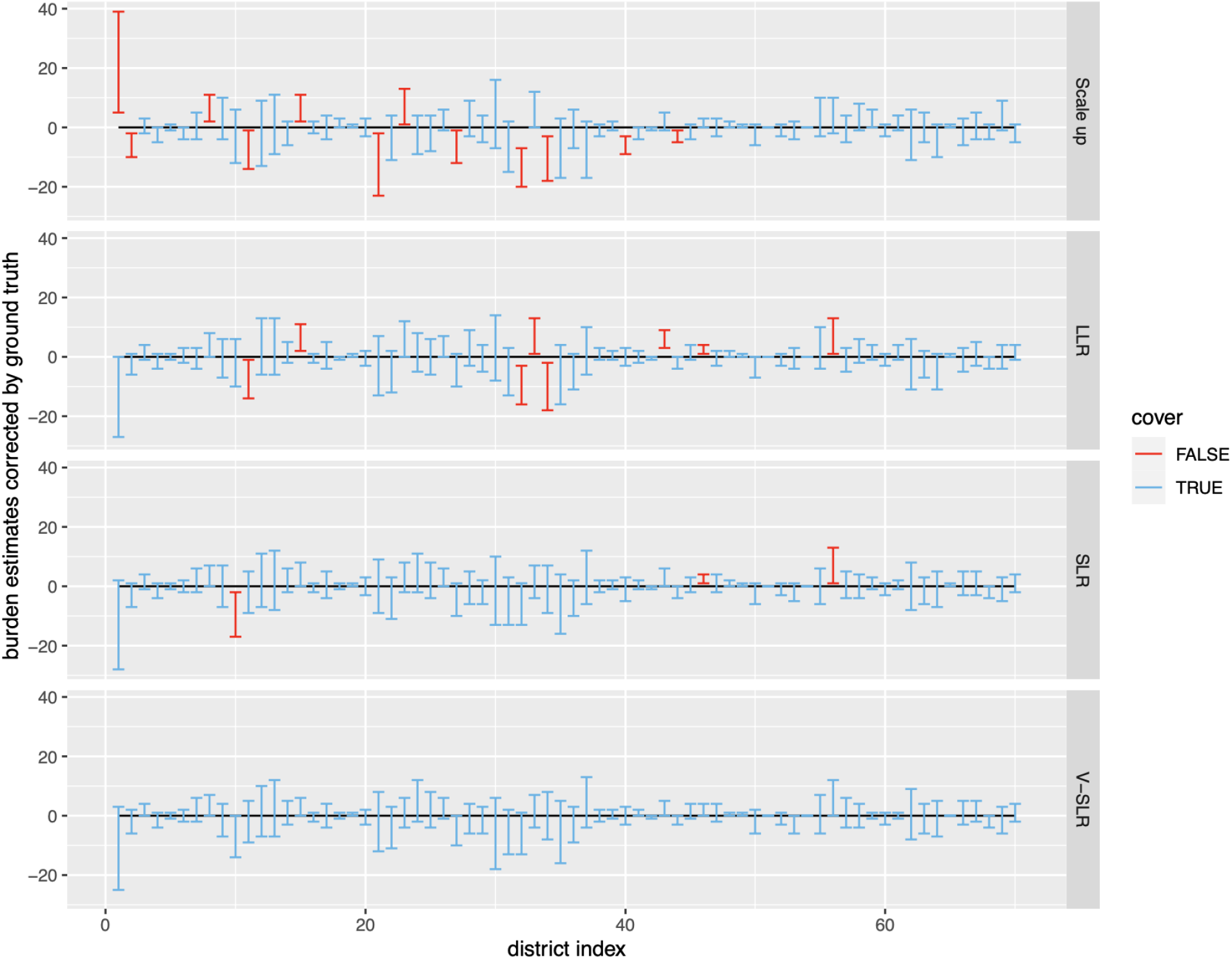
95% confidence interval for burden estimates of scaling method, LLR, SLR and V-SLR model on district level.

### 5.3 Burden Estimation on Suspected Cases

After comparing the performance of scaling up method, LLR, SLR and V-SLR on both classification and burden estimation, we choose and apply V-SLR to conduct further study on providing a model-based burden estimation on the 31,853 suspected reported cases without laboratory confirmation. To investigate the influence of using symptomatic case definition, we study the relationship between inferred measles burden and the number of reported suspected cases and create a descriptive statistic, ratio between the number of predicted positives and the reported suspected cases. Figure 6 (a) shows the fluctuation of this ratio (with 95% confidence interval bands) from 2009 to 2017 by month and is aligned with the number of all suspected cases to better show the trend in time. For time intervals with high suspected cases, the ratio tends to go lower and when the number of suspected cases is low, the ratio is higher. It indicates that the ratio between (inferred) measles cases and the reported febrile and rash cases is not constant across time. We note that the assumption of constant disease rate among the reported cases over time is widely used in burden estimation methods based on time series studies without taking individual-level demographic and geolocational effects into account. When we aggregate the plot by year, figure 6 (b) shows a more stable ratio across time regardless of the movement of the number of reported cases. By comparing the two plots, though we observe a stable disease rate by year, there is fluctuation in this rate in finer time units which may due to the heterogeneity of people in various aspects such as age, vaccination status, location and time of visit.

**Figure 6:**
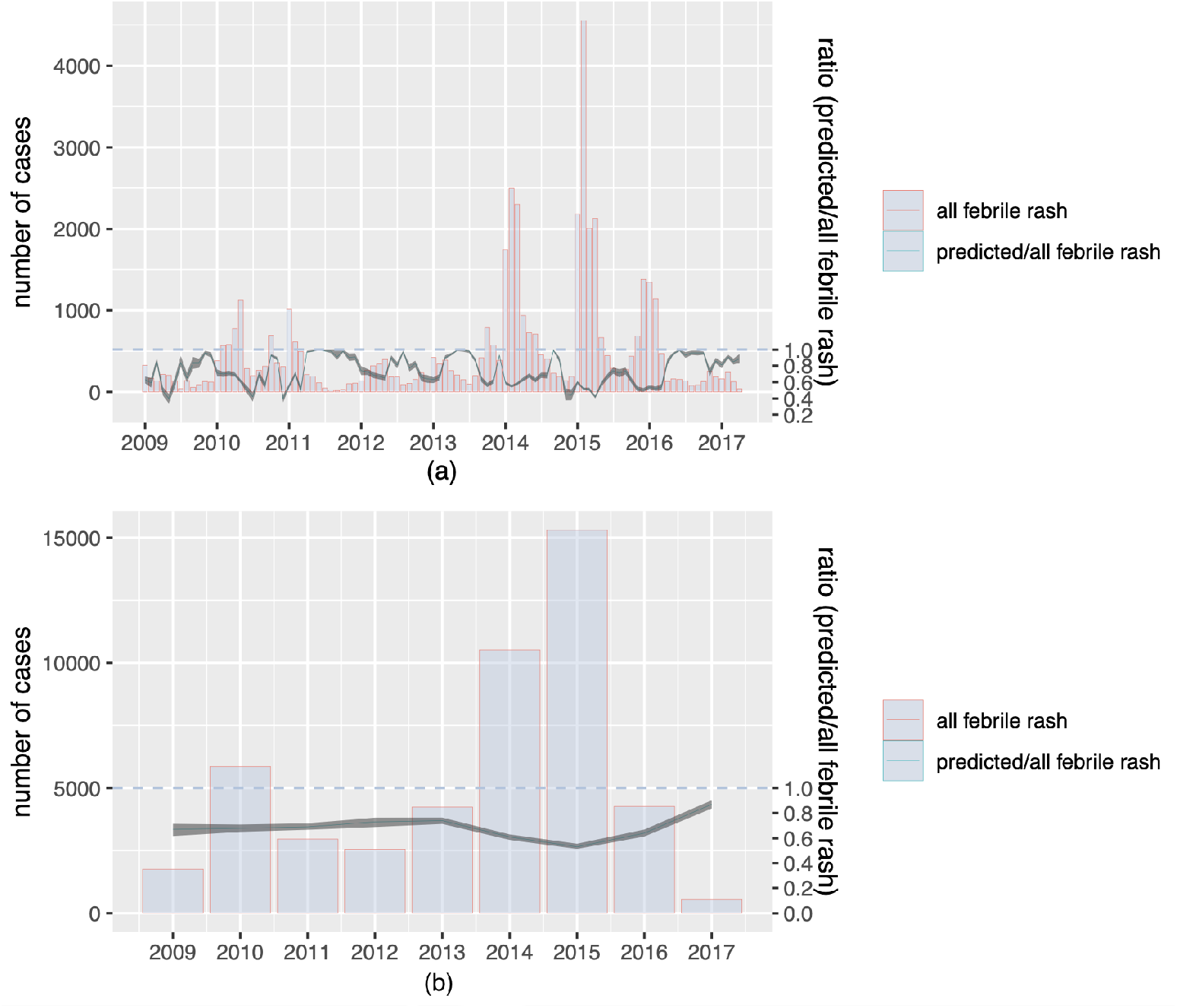
Ratio between predicted and suspected cases aligned with number of suspected cases, aggregated by (a) month and (b) year

## 6 Conclusion and Discussion

We have proposed a spatially varying coefficient model (V-SLR) that allows to integrate both clinically confirmed and diagnostically confirmed measles cases to generate a better estimate of the burden of reported measles at district-level in Ethiopia. We compared the performance of the proposed model with a naive scaling up method and two other useful approaches that we called LLR and SLR, both in terms of individual-level classification and burden estimation on reported suspected cases. Both V-SLR and SLR can incorporate some level of low quality data issues such as missingness and imbalance by utilizing information from neighbors through spatial dependence. In the simulation study and real data study, all the methods can provide a reasonable confidence interval of burden estimation at country level, while the proposed method outperforms the other methods in confidence intervals at district level, which provides more accurate indication about the measles epidemic with spatial heterogeneity taken into account. We apply our proposed method to the suspected reported cases with febrile rash and bring out some insights on the influence of using symptomatic cases definition on monitoring measles disease burden. Naively, relying on uncorrected, clinically confirmed, suspected cases over-report the burden of reported cases. More importantly, the magnitude of that bias is non-constant in time and varies as a function of the location, age-distribution, and vaccine history of cases presenting with febrile rash. This latter point is important for national-level burden estimation and vaccine program evaluation as the conventional state-space models that are fit to infer these quantities conventionally assume that reporting rates are constant in time (Dixon et al., 2021; Thakkar et al., 2019). We note here that these results only address time varying diagnostic uncertainty due to variability in the predictive value of the clinical case definition and do not address temporal or spatial variation in care-seeking.

Here we have presented a proof of concept for the use of diagnostic testing results to develop a classifier and infer burden of infection among untested, clinically confirmed cases. We note that, in doing so, we have made the strong assumption that tested cases are a representative sample of clinically compatible cases. In practice, this is unlikely to be the case, either for this data set, or for applications in other countries, as there is not formal guidance on the application of diagnostic testing to ensure a representative sample. Thus, the sample of tested individuals here is a convenience sample and may therefore result in unforeseen biases. We note that while there are consistent patterns in the fitted parameters (e.g. individuals with a prior vaccination had lower odds of being measles IgM positive), interpretation of these parameters must acknowledge the potential for bias due to non-random sampling. The substantial gap between assumption and reality could be overcome with a formal sampling design, intended to take advantage of the methods we describe above. Our individual-level classifier and inference about measles burden provide a way to utilize the current surveillance data and reform them into a representative sampling and testing diagnostic, which provide an entrance to analyze the long-lasting chicken-and-egg problem: the co-dependence between the formation of hard-to-achieve perfect surveillance data and the development of methodology to analyze surveillance data better.

## Data Availability

These data are the property of the government of Ethiopia and the WHO. Access to these data can be requested directly from WHO Division of Immunization, Vaccinations, and Biologicals. To illustrate the application of the methods described herein, we have provided the readers with a simulated data set used in our manuscript. These data have similar characteristics to the real data: https://github.com/lxxiww/Measles-Surveillance

## Footnotes

1 Ethiopia is administratively divided into four levels: regions (provinces), zones, woredas (districts) and kebele (wards)(Wubneh, 2017).

2 distances are measured in km

