## Supplementary Material for "Measles Burden Estimation Using Local Gaussian Process Classifiers"

### Epidemiology and Infection: Measles Burden Estimation Using Local Gaussian Process Classifiers

#### Supplementary Material

For prior specification of unknown parameters, in V-SLR, we give inverse gamma prior with shape parameter 0.2 and scale parameter 0.2 on $\sigma^2_{\beta_k}$ and $\sigma^2_w$, denoted as $\sigma^2_{\beta_k} \sim$ InverseGamma$(0.2, 0.2)$ and $\sigma^2_w \sim$ InverseGamma$(0.2, 0.2)$. The prior for $\phi$ is uniform and lower bound 0 and upper bound depends on the map of consideration. For the mean correcting parameter $\epsilon$, we impose a normal prior with mean 0 variance 1. In LLR, we impose normal prior on the $\beta$'s for each predictor at each location, i.e., $\beta_k(s) \sim \mathcal{N}(0, 4)$ to allow for more flexibility in term of coefficient values. In SLR, we adopt the Normal prior on $\beta$'s as in LLR and inverse Gamma with shape parameter 0.2 and scale parameter 0.2 on the variance parameter $\sigma^2$. We provide the In the next subsection, we introduce the details of model fitting using Markov Chain Monte Carlo (MCMC).

##### Markov Chain Monte Carlo (MCMC) Algorithm

We use Markov chain Monte Carlo (MCMC) for carrying out Bayesian inference for the LLR, SLR and V-SLR models. For efficiency and extendability, we implement the MCMC algorithms using the language Nimble (de Valpine et al., 2017). In this section, we provide the details of MCMC algorithm for model fitting.

In LLR, the prior of the coefficient for each predictor $k$ at location $s$ follows a Normal distribution independently, $\beta_k(s) \sim \mathcal{N}(0, 4)$, and the prior for the intercept is $\beta_0(s) \sim \mathcal{N}(0, 4)$. The full conditional distribution for $\beta_k(s)$ with $k \in \{0, ..., p\}$ is then

*

$$\pi(\beta_k(s)|.) \propto \prod_{i=1}^{J_s} f(Y_i(s)|\beta_0(s), ..., \beta_p(s))p(\beta_k(s))$$

where $f(Y_i(S)|.) = P(Y_i(s)|.)^{Y_i(s)}[1 - P(Y_i(s)|.)]^{1-Y_i(s)}$ and $P(Y_i(s)|.) = \frac{1}{1+\exp(-\boldsymbol{X}_i(s)\boldsymbol{\beta(s)}-\beta_0)}$. Since no spatial dependence is assumed between locations, LLR can be easily parallelized in computation and is most computational efficient among the three methods.

In SLR, spatial dependence is conveyed through modeling the random effects using Gaussian process with mean 0 and covariance function that decrease with distance, i.e., $W \sim GP(0, \Sigma(\sigma^2, \phi))$. The prior for $\phi$ is uniform with lower bound 0 and upper bound depending on the range of map under consideration. Prior for $\sigma^2$ follows an inverse gamma prior specified in Section 3.1. The posterior distribution for $W(s)$ is

$$\pi(\boldsymbol{W}|.) \propto \prod_{s\in\mathcal{D}} \prod_{i=1}^{Js} f(Y_i(s)|W(s), \beta_1, ..., \beta_p, \sigma^2, \phi)p(\boldsymbol{W}|\sigma^2, \phi)p(\sigma^2)p(\phi)$$

. The Gaussian process samples from $p(\boldsymbol{W}|\sigma^2, \phi)$ are realized through a Metropolis-Hastings adaptive random-walk sampler with a multivariate normal proposal (Roberts and Sahu, 1997). As for the coefficient of the fixed effects $\beta_k$ , the posterior is as follows

$$\pi(\beta_k|.) \propto \prod_{s\in\mathcal{D}} \prod_{i=1}^{J_s} f(Y_i(s)|W(s), \beta_1, ..., \beta_p, \sigma^2, \phi)p(\beta_k)$$

where $k \in \{1, ..., p\}$ among the $p$ predictors. The hyperparameters for the covariance matrix are also updated as the fixed effects.

In V-SLR, we further describe the spatial dependency through $p+1$ Gaussian processes on both covariates and random effects. It is reasonable to assume same spatial scaling parameter $\phi$ shared across these Gaussian processes since the spatial dependence is all on one map, and is also supported by empirical experiences that the estimation for $\phi$ is usually unstable compared to other spatial parameters. Let $\Theta = \{\phi, \sigma^2_{\beta_1}, ..., \sigma^2_{\beta_p}, \sigma^2_W\}$ be the set for hyperparameters of $\Sigma_{\beta_K}$ and $\Sigma_W$. The posteriors for the covariates and random effects are:

$$\pi(\boldsymbol{\beta}_k|.) \propto \prod_{s\in\mathcal{D}} \prod_{i=1}^{Js} f(Y_i(s)|W(s), \beta_1(s), ..., \beta_p(s), \Theta, \epsilon)p(\boldsymbol{\beta}_k|\sigma^2_{\beta_k}, \phi)p(\sigma^2_{\beta_k})p(\phi)$$

$$\pi(\boldsymbol{W}|.) \propto \prod_{s\in\mathcal{D}} \prod_{i=1}^{Js} f(Y_i(s)|W(s), \beta_1(s), ..., \beta_p(s), \Theta, \epsilon)p(\boldsymbol{W}|\sigma^2_w, \phi)p(\sigma^2_w)p(\phi)$$

Same as SLR, we also use Metropolis-Hastings adaptive random-walk sampler to generate samples from $p(\boldsymbol{\beta}_k|\theta_{\beta_k})$ and $p(\boldsymbol{W}|\theta_w)$. As for the mean correcting parameter $\epsilon$, we assign $\epsilon \sim \text{Uniform}(-10, 10)$ as prior information and the posterior is as follows,

$$\pi(\epsilon|.) \propto \prod_{s\in\mathcal{D}} \prod_{i=1}^{Js} f(Y_i(s)|W(s), \beta_1(s), ..., \beta_p(s).\Theta, \epsilon)p(\epsilon)$$

Hyperparameters in $\Theta$ are updated using conditional density functions obtained using the same step as $\epsilon$.
